# Assessing the Mental Health Crisis Among New York’s Postdoctoral Researchers

**DOI:** 10.1101/2025.06.24.25328454

**Authors:** Silvia Martinelli, Jazib Uddin, Estibaliz Barrio Alonso, Raura Doreste, Sahar Jalal

## Abstract

Grave concerns have been raised in recent years regarding the graduate student mental health crisis in academia; however, similar attention has not been focused on the subsequent and more challenging career stage in science, that of the postdoctoral researchers. We conducted a pilot survey among postdoctoral associates (*N* =160) at Weill Cornell Medicine, Cornell University’s medical school in New York City, to understand the unique challenges faced by the postdoctoral community. Our survey found that respondents’ mental health and wellness were primarily affected by challenges intrinsic to the demands of academic life, with additional factors related to living in a major metropolitan city. Additionally, the survey identified unique challenges of postdoctoral researchers based on their gender, ethnicity, or citizenship/immigration status. Finally, our analyses also found that respondents had a negative outlook on career progression and ability to transition into an independent position within academia. These results are in line with similar surveys conducted in the past among students and postdoctoral researchers, and highlight the fact that, despite critical evidence, the situation has not changed overtime. Further studies should be conducted across various institutions and cities to gather more comprehensive data on the mental health and wellbeing of early-stage academic researchers. The findings from this study can be utilized by institutional Postdoctoral Offices or Associations to provide updated policies and resources aimed at alleviating some of the professional stresses faced by postdoctoral researchers.

## INTRODUCTION

Postdoctoral fellowship is an ill-defined stage in the path of a typical academic research career. Research training is the critical process of teaching individuals the skills, methods, and techniques needed to conduct scholarly research^1^. It typically involves instruction in research design, data collection, data analysis, critical thinking, and academic writing. Postdoctoral training refers to the non-permanent training period after one has procured their doctoral degree and before securing a more long-term faculty or industry position, where the experiences gained during these years facilitate the transition to a career as an independent scientist^2^. In an ideal system, postdoctoral researchers gain critical research experience alongside valuable career guidance from an accomplished mentor who has successfully navigated the same journey beforehand. Importantly, during this period postdoctoral researchers learn to develop ideas for their independent research, apply for fellowships and grants, develop their own professional networks, and publish their novel work^3^. Traditionally, accomplishing these benchmarks would allow for postdoctoral researchers to move into tenure-track research faculty positions. However, as the number of postdoctoral researchers continues to expand, the number of tenure-track job openings has not grown at a similar pace to meet increased demand^4^. Furthermore, strict time limitations on postdoctoral positions, typically capped at five years, place significant pressure on researchers to secure permanent positions within an unrealistic timeframe. Compounding this issue is the lack of structured career development and support systems for postdoctoral researchers to transition into alternative nonacademic career paths, leaving many without clear pathways or incentives to pursue careers outside academia^5^. These challenges create a bottleneck where postdoctoral researchers find themselves unable to advance to either academic faculty or industry positions. Consequently, this lack of viable long-term career prospects deters graduate students from pursuing postdoctoral fellowships altogether and has culminated in the current shortage wherein academic institutions are reporting challenges in recruiting postdoctoral researchers^6–8^.

Postdoctoral researchers are a vulnerable group in academia due to the pressure to succeed alongside growing professional and personal commitments. The duration of time spent in this position varies greatly, both across and within fields, as does the age range of postdoctoral fellows. This variability reflects differences in life stages, including family formation and broader personal circumstances. At work they find themselves between being highly skilled experts in a specialized area yet lacking broader career experience. This transition phase is paralleled legally by often-ambiguous employment contracts—postdoctoral researchers are no longer students but are also not fully defined as employees, leading to complex legal and financial implications. Correlated issues include low pay compared to the cost of living and equivalent-stage workers ^9,10^, mistreatment by supervisors and systems that triggers chronic stress and burnout, and uncertainties regarding unstable visa status in the case of international researchers^10,11^. Additionally, postdoctoral researchers suffer from a persistent sense of insecurity caused by the pressure to leave the academic pipeline if the publishing record is inadequate or due to limitations with acquiring independent grant funding^12^. Overall, several studies have shown that postdoctoral researchers have severe work-life imbalance issues, which further impact their mental health negatively^13–15^.

Importantly, having adequate mental health is essential for producing quality scientific work. Studies have shown that medical residents struggle to reconcile their work with their personal lives ^16,17^. While research postdoctoral researchers also struggle with these challenges, similar attention has not been focused on the researchers, as there are not many studies that have examined these concerns^18^. One exception is a study showing how the COVID-19 pandemic had impacted the mental health of postdoctoral associates^19^. As an example, travel restrictions imposed because of the pandemic exacerbated feelings of loneliness and isolation and negatively impacted pressures to succeed during the postdoctoral training period. Hence, it is important to know more about the current stressors among postdoctoral trainees and examine how these are detrimental factors for their mental health. It is essential for postdoctoral researchers to accomplish their research in supportive environments, highlighting the need of institutions to prioritize their mental health. Thus, we sought to measure the extent of mental health struggles among postdoctoral researchers in a major research institution based in a metropolitan city.

The set of challenges that postdoctoral researchers face extends beyond the academic pressures of their job roles, particularly when living in a major metropolitan city. The high cost of living severely strains budgets, making it difficult to find affordable housing while receiving a salary that is, with few exceptions, below the state average salary^9^. Access to affordable childcare is another significant concern; long hours in the lab can complicate finding reliable, nearby options, leaving many researcher-parents feeling torn between their professional and familial responsibilities^20^. Counterintuitively, living in a major city can contribute to feelings of isolation and loneliness, as the demanding work schedules make it hard to build supportive networks outside of academia^21^. These struggles can profoundly impact both well-being and productivity, highlighting the need to better investigate the unique struggles for postdoctoral researchers so that we can build better support systems from them within the research community.

Considering these factors, we aimed to gather survey data of the postdoctoral community at Weill Cornell Medicine (WCM) in New York City, as a case study to understand the unique perspectives and burdens of postdoctoral researchers living in a major metropolitan city. Our study first assessed the general mental health and well-being of survey respondents and identified factors that are intrinsic to the postdoctoral community at WCM. Our survey also considered the demographics of our respondents (i.e., gender, citizenship, and ethnicity) and how those identities informed the unique challenges that are experienced by different members of the community. Further, we examined how current resources available at WCM to support the well-being of postdoctoral researchers are utilized and whether they are currently adequate. Finally, we examined how the COVID-19 pandemic has affected the mental health of WCM postdoctoral researchers. Altogether, our survey results provide insights into the mental health and wellbeing of a group of postdoctoral researchers at a unique career stage in academic research that has often been ignored.

## RESULTS

### Description of the Postdoctoral Mental Health Survey

This study utilized an observational exploratory survey to evaluate the wellbeing and overall satisfaction of postdoctoral researchers at WCM and to identify new areas for intervention that can improve the mental health status of postdoctoral researchers. The anonymous survey results aim to guide Postdoctoral Associations in shaping future initiatives and informing discussions with the Office of Postdoctoral Affairs and institutional leadership. By examining the mental health status of current postdoctoral researchers, the study seeks to identify critical areas of need, including symptoms of depression, anxiety, and burnout, while considering the additional impacts of the COVID-19 pandemic. The survey also explores job-related stressors, such as workplace challenges and experiences of aggression, to gain a comprehensive understanding of factors affecting postdoctoral life. The primary objective is to uncover key stressors and highlight areas where institutional support is effective, as well as areas requiring improvement. The survey incorporates questions adapted from established tools like the PHQ-9 and GAD-7 to assess mood-related symptoms, alongside custom-developed questions designed to evaluate other dimensions of wellbeing, such as cognitive function and social engagement^22,23^. These de novo items, partially informed by earlier pilot testing in 2021, aim to capture unique stressors and experiences specific to the WCM postdoctoral community.

### Demographic Information of Survey Respondents

A total of 198 postdoctoral associates at WCM participated in the survey conducted between December 2023 and January 2024 by the Weill Cornell Postdoctoral Association (PDA). Of these, 160 finished the survey and gave their consent to share the outcome. Of these respondents, 40.5% (65) self-identified as male, 55% (88) as female, 4% (6) as non-binary, and 0.5% (1) preferred not to say (**Figure 1A**). We also found that 74% (99) of the responders were nonimmigrant work visa-holders, 7% (9) were permanent residents (green card holders), and 19% (25) responders were US citizens (**Figure 1B**). For the purpose of this study, we also asked respondents whether they belong to an ethnic minority population that is underrepresented in the medical profession relative to their numbers in the general population (UriM, Underrepresented in Medicine) or are of Asian ethnicity, a minority group that is not considered underrepresented in medicine^24^. In our survey, 11% (18) responders self-identified as UriM whereas 29.5% (47) identified as Asian; the remaining 59.5% (95) responders did not identify as belonging to either of these groups and were grouped into “Other Ethnicity/Undisclosed” (**Figure 1C**).

**Figure 1.**
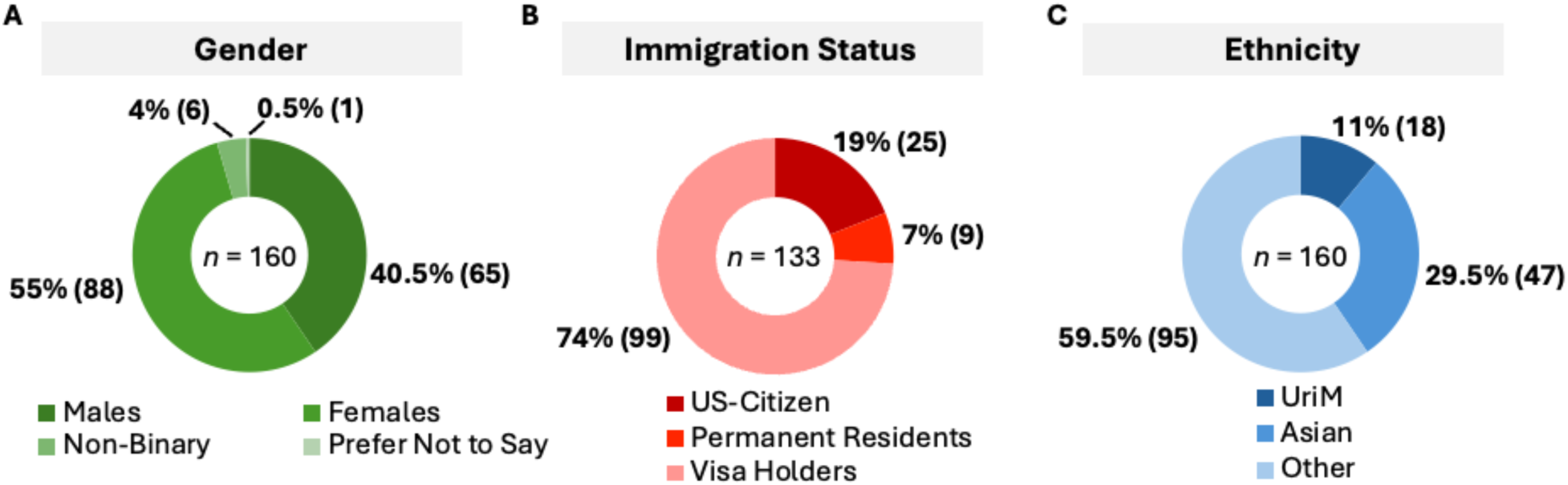
Demographics of the surveyed population stratified by **A**) gender, **B**) immigration status and **C**) ethnicity; *N*=160. The total number of respondents for the Immigration Status is below 160 due to omitted responses.

### Stress-related Symptoms of Postdoctoral Researchers

The respondents were asked to answer whether and how often in the previous 30 days they experienced symptoms that are often associated with severe stress and depression. Of the 160 respondents, 48.13% (77) reported always or frequently having trouble relaxing, 46.25 % (74) (feeling overwhelmed by responsibilities, 45.63% (73) feeling nervous, and 44.38%(71) being depleted of energy. Additionally, between 30% and 40% of respondents indicated they frequently or always experienced various anxiety-like symptoms affecting their social motivation and sleep quality [Feeling down or sad, 39.38% (63); Having a recurrent feeling that something negative might happen, 35.63% (57); Little interest in socializing, 35.01% (56); Sleeping issues, 35% (56); Trouble concentrating, 32.5% (52); Fear of having disappointed important people in life, 32.5% (52)]. More than one in four also reported mood impacts, such as increased irritability (27.5%, 44), as well as stress-related changes in eating habits (27.5%, 44) and slower thinking processes (26.88%, 43). Stronger physical effects, such as body disconnection and changes in movement and speech, were also reported as always or frequently, but by a smaller percentage [16.88% (27) and 10.01% (16) respectively) **(Table 1 and Table S1)**.

**Table 1.** Percentages of responses to the question “In the last 30 days, how often have you felt the following?” with possibility of single choice selection between “never”, ”rarely”, “sometimes”, frequently”, “always”. The table shows summed percentages for categories “never” and “rarely”, as well as “frequently” and “always”. *n* = 160

| Feeling/Mood | Never/Rarely | Sometimes | Frequently/Always |
| --- | --- | --- | --- |
| Trouble relaxing. | 15.63% | 36.25% | 48.13% |
| Feeling overwhelmed by responsibilities. | 20.63% | 33.13% | 46.25% |
| Feeling nervous. | 15.00% | 39.38% | 45.63% |
| Being depleted of energy. | 22.50% | 33.13% | 44.38% |
| Reduced self confidence. | 25.00% | 35.00% | 40.00% |
| Feeling down or sad. | 20.63% | 40.00% | 39.38% |
| Having a recurring feeling that something negative might happen. | 32.51% | 31.87% | 35.63% |
| Little interest in socializing. | 31.25% | 33.75% | 35.01% |
| Sleeping issues. | 33.13% | 31.87% | 35.00% |
| Trouble concentrating. | 29.38% | 38.13% | 32.50% |
| Fear of having disappointed important people in my life. | 39.38% | 28.13% | 32.50% |
| Becoming easily annoyed or irritable. | 33.13% | 39.38% | 27.50% |
| Poor appetite or overeating. | 40.00% | 32.50% | 27.50% |
| Slowed thinking process. | 36.25% | 36.88% | 26.88% |
| Feeling disconnected from your body. | 66.26% | 16.88% | 16.88% |
| Noticeable changes in movement speed or speech rate. | 73.75% | 16.25% | 10.01% |

### Work-related Parameters Affecting Mental Health

To evaluate potential work-related factors contributing to self-reported stress symptoms, respondents were asked to identify various parameters related to both intrinsic aspects of the work environment and external, work-dependent influences affecting their mental health and overall wellbeing. Strikingly, salary was reported as the leading factor affecting the mental health and wellbeing of postdoctoral researchers. Salary remains the main cause of distress across various stratifications - gender, ethnicity, or immigration status - being selected by more than 80% (136) of respondents within each group, underscoring the robustness of this finding, with even increased percentages of more than 85% (93) when considering only international researchers (**Figure 2A-D).** The only exceptions are the non-binary/gender non-conforming and undisclosed categories, which can be attributed to the small sample size (7 respondents, of which 5 indicated salary as a factor affecting mental health) (**Figure 2B**). Current WCM salary policy dictates that incoming postdoctoral researchers should be offered a minimum yearly gross salary of $61,008, or monthly gross salary of $5,084, with annual increases based on experience^25^. However, this monthly salary falls below the estimated basic living costs of $5,830 per month in North Manhattan, excluding childcare^26^. This disparity is consistent among larger cities and suggests why salary was indicated as of the main distresses for work related parameters that affect postdoctoral mental health^9^. Furthermore, among all postdoctoral researchers and in any subgroup, more than half the respondents indicated that work/life balance (67%), the housing situation (56%) and job security (51%) heavily affected their wellbeing (**Figure 2A**). Stratification by gender revealed an overall equal distribution between females and males of perceived stressors with few exceptions: housing appears to be a higher stress factor for males (male: 65% vs female: 50%), whereas access to mentoring (female: 32% vs male: 21%) and healthcare assistance (female: 21% vs male: 15%) seem to be a higher stress burden for women (**Figure 2B**). Intuitively, visa/immigration issues, healthcare assistance, and job security represent a stronger stress factor among visa holders relative to US-citizens and permanent residents (63% vs 6%, 24% vs 6%, and 59% vs 30% respectively; **Figure 2D and Figure S1**). On a positive note, discrimination is among the lower factors affecting mental health, even when stratifying for UriM and Asian ethnicities (identified as stressor by 11% and 13% respectively, and 10% overall) (**Figure 2A and C**).

**Figure 2.**
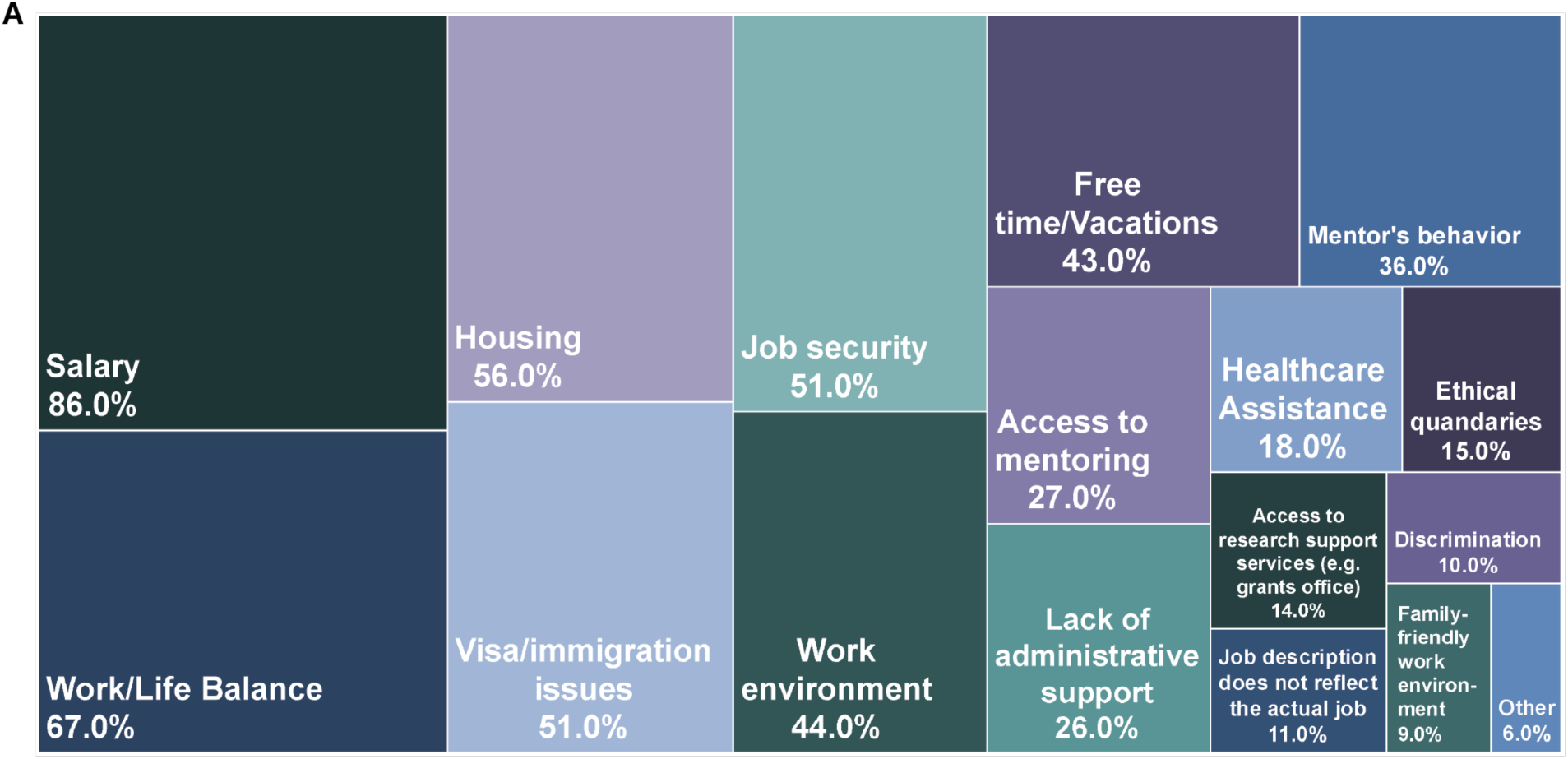
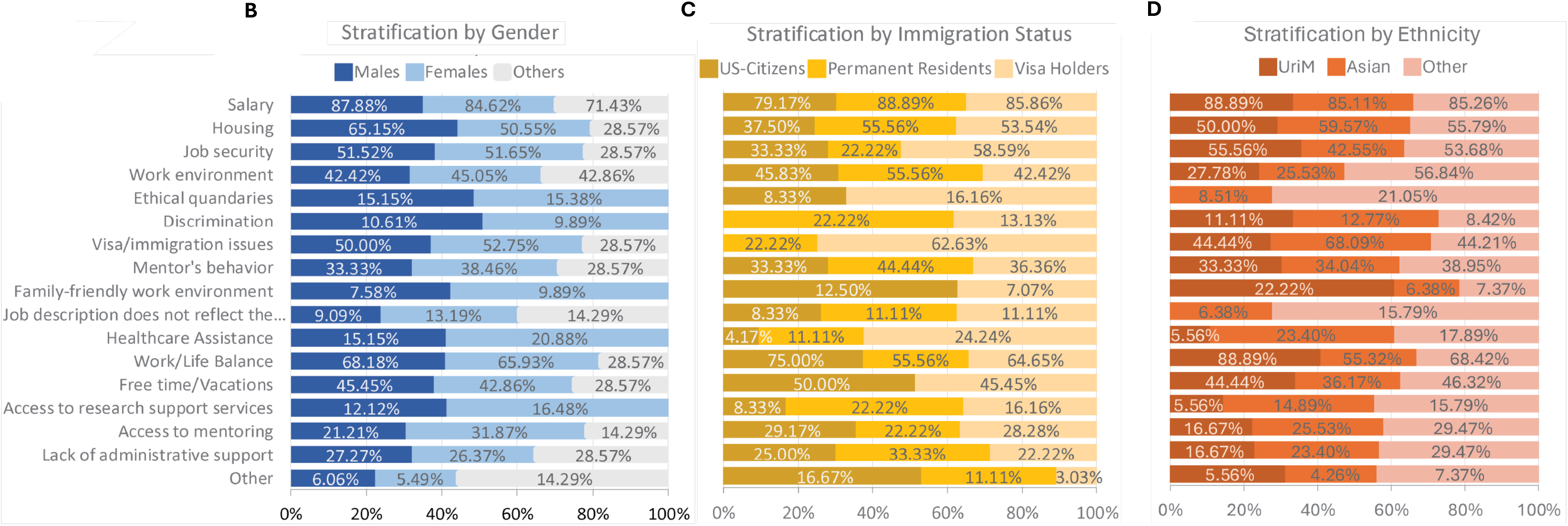
Work-related factors that impact postdocs’ mental health and general wellbeing. **A**) Chart representing which percentage each factor was selected by the all the respondents (*n* = 159). **B**) Stratification of A based o gender. **C**) Stratifciation of A based on ethnicity. **D**) Stratification of A based on Immigration status.

To gain a more detailed understanding of work-specific stressors, postdoctoral researchers were additionally asked to identify whether any common work-related conflicts or challenges were regularly affecting their mental health. This step aimed to deepen the insights gained from the previous questions by focusing specifically on workplace dynamics. Overall, 52.5% of all respondents answered “yes” and 24.4% of respondents answered “sometimes” when asked whether the pressure to work at optimum levels all the time affected their mental health (**Table 2 and Table S2**). An interesting divergence was observed when looking at groups that believe that job performance pressure does not affect their mental health and well-being; 31% of males and 34% of Asians answered “no” when they when asked whether the pressure to work at optimum levels all the time affected their mental health, while only 17% of women, 18% of people belonging to UriM and 19% of people belonging to other ethnic groups had the same answer. This suggests that high performance stress on the workplace affects less males and Asians. All the proposed situations, except for insufficient support for dependents, were selected by more than half of the 160 respondents as regularly affecting their mental health at least sometimes (**Table 2 and Table S2**), emphasizing the pervasive nature of intrinsic stress that continuously accompanies the daily work life of postdoctoral researchers.

**Table 2.** Percentages of responses to the question “ In the last 30 days, how often have you felt the following?” with possibility of single choice selection between “never”, ”rarely”, “sometimes”, frequently”, “always”. The table shows summed percentages for categories “never” and “rarely”, as well as “frequently” and “always”. *n* = 160

| Work-related Stressor | Yes or Sometimes | No | Total (n) |
| --- | --- | --- | --- |
| Pressure to work at optimum levels, all the time | 76.90% | 23.10% | 160 |
| Lack of career development opportunities | 75.60% | 24.40% | 160 |
| Relationship with PI | 68.70% | 31.30% | 160 |
| Dependency between performance and our visa status | 56.90% | 43.10% | 160 |
| Relationship with co-workers | 55.10% | 45.00% | 160 |
| Lack of clarity regarding vacation policy | 50.00% | 50.00% | 160 |
| Insufficient support for dependents | 43.80% | 56.30% | 160 |

### The Influence of Micro- and Macro-aggressions on Mental Health

To assess experiences of bias and exclusion, postdoctoral researchers were first provided with the definitions of microaggressions as everyday slights, indignities, put-downs, and insults directed at people of color, women, LGBTQ+ individuals, and other marginalized groups in daily interactions^27^, and macroaggressions as the beliefs and ideologies that reinforce microaggressions and institutionalized racism^28^. They were then asked whether they had experienced micro- or macroaggressions within WCM and whether these experiences had impacted their mental health. 57% (91) of the responders reported to have experienced microaggressions. Of these, 40% (36) reported that it has not much affected their mental health, 25% (23) felt little effect, another 25% (23) moderate effect, and only 10% (9) experienced an extensive effect (**Figure 3A**). Macroaggressions, on the other hand, appeared to have a slightly lower impact, affecting roughly half of the respondents (49%, 82), of which 43% (34) being not much affected, 24% (19) a little, 27% (20) moderately and only 6% (5) extensively (**Figure 3A**). Since micro- and macro-aggressions are often affecting more minorities or marginalized groups, we looked at the population of the respondents who were more affected (extensively and moderately) by either micro- or macro-aggressions. Of the 44 respondents affected, the majority belonged to non-US working visa holders (34%, 15), while only one green-card holder (2.2%, 1) and no US citizen (0%, 0) were part of this group (**Figure 3B**). The distribution between females (20.5%, 9) and males (27.2%, 12) was similar, and among the racial groups, only Asians appeared in this group with 16% (7) of the total. None of the persons identifying as non-binary (0%, 0) nor UriM (0%, 0) reported to be moderately or extensively affected by micro- and macro-aggressions, representing a positive surprise given the high incidence of such aggressions among minorities and underrepresented groups (**Figure 3B**). Confirming the self-report on experiencing micro- and macro-aggression, when asked whether they had observed such aggressions impacting other people, over 51% of the respondents answered yes, 40% did not notice and only 8.8% answered no (**Figure 3C**).

**Figure 3.**
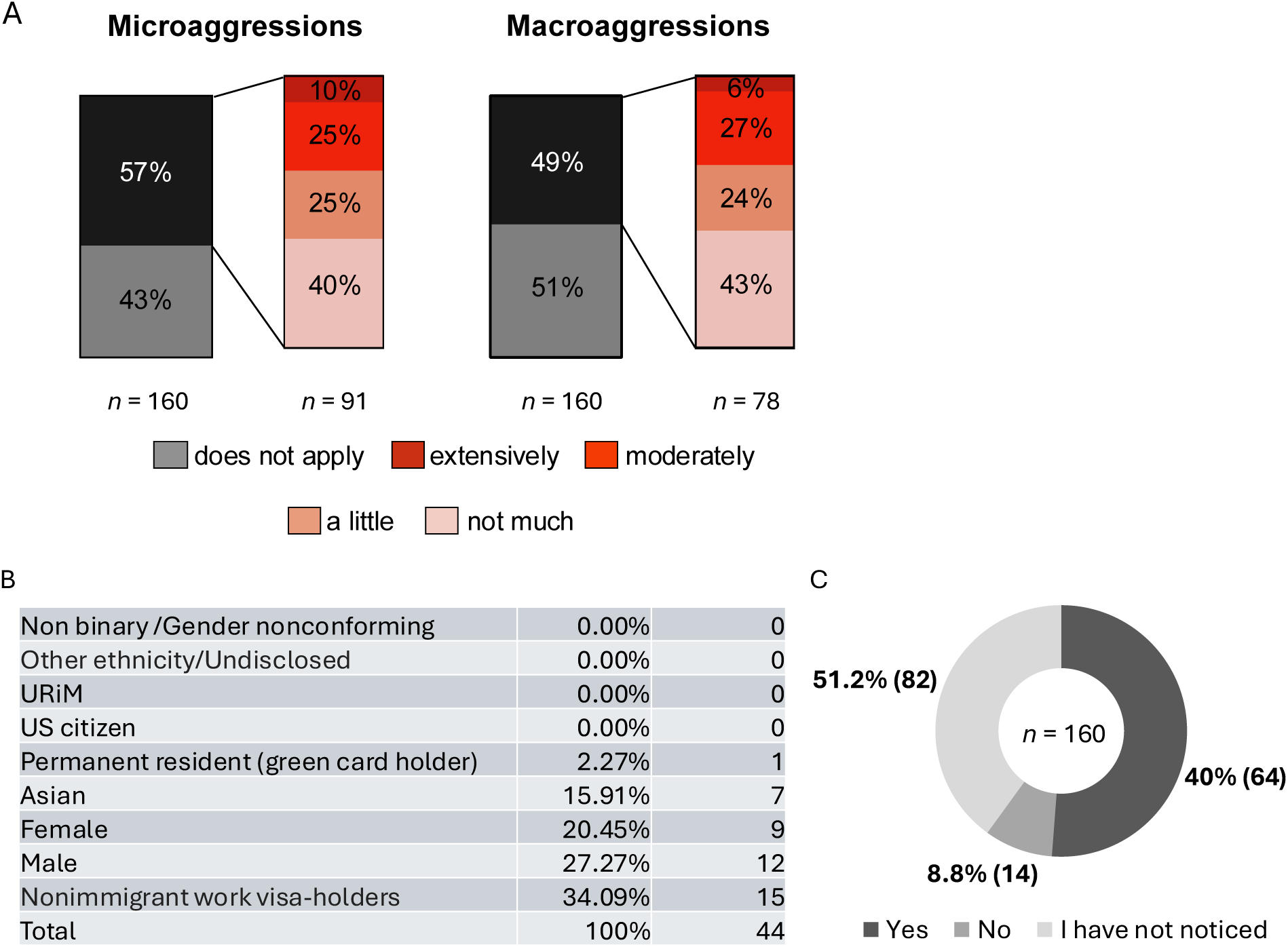
**A**) The impact of micro- and macro-aggressions on mental health, if experienced. The categories "Not Much," "A Little," "Moderately," and "Extensively" represent increasing levels of severity. "Does Not Apply" indicates the absence of any experience with micro- and macro-aggressions. **B**) Stratification by gender, race and immigration status of respondents that were affected moderately or extensively by micro- or macroaggressions. **C**) Percentages of responses to the question “If not personally affected, do you have the perception that people suffer from macro/microaggression at WCMC?”, *n*=160.

#### The long-term impact of the pandemic on mental health

Given the tremendous impact the COVID-19 pandemic has had worldwide on all aspects of our lives, including the mental health of academic researcher^29^, we wanted to investigate whether and to what extent it might still contribute to postdoctoral researchers’ mental health at the end of 2023. The first question inquired the perception of the COVID-19 pandemic’s risk to the postdoctoral researchers’ mental health on an increasing scale from “no risk at all”, “minor risk”, “significant risk” up to “major risk”, with additional possibilities of “don’t know” and “prefer not to answer”. The data revealed a generally low perception of risk, with nearly half of respondents identifying it as a minor risk (46%) (**Figure 4A**). Nevertheless, 20% regarded it as significant risk, and 5% as a major risk. Notably, these percentage were lower in certain subgroups: only 15% of males and gender non-conform people perceived the pandemic as a significant risk (versus 21% overall and 24% females), and 1.5% of males as a major risk (versus 5% overall, 8% females and 14% gender non-conform). Furthermore, none of the U.S. citizens and permanent residents surveyed considered the pandemic a major risk; while among Asians, the perception of the pandemic as a major risk was less than half of the overall average, with a 2.1% selection by the respondents (**Supplementary Data**). Almost one in five respondents (18.8%) regarded the pandemic as having no risk at all on their mental health (a similar percentage of the population viewing the pandemic as a significant risk, 20.6%), while 8.8% reported “don’t know” and 1.3% preferred not to answer (**Figure 4A**). Overall, the results show a gaussian distribution of responses around the middle value “minor risk”, indicating that the pandemic had (luckily) significantly lowered its impact by end of 2023.

**Figure 4.**
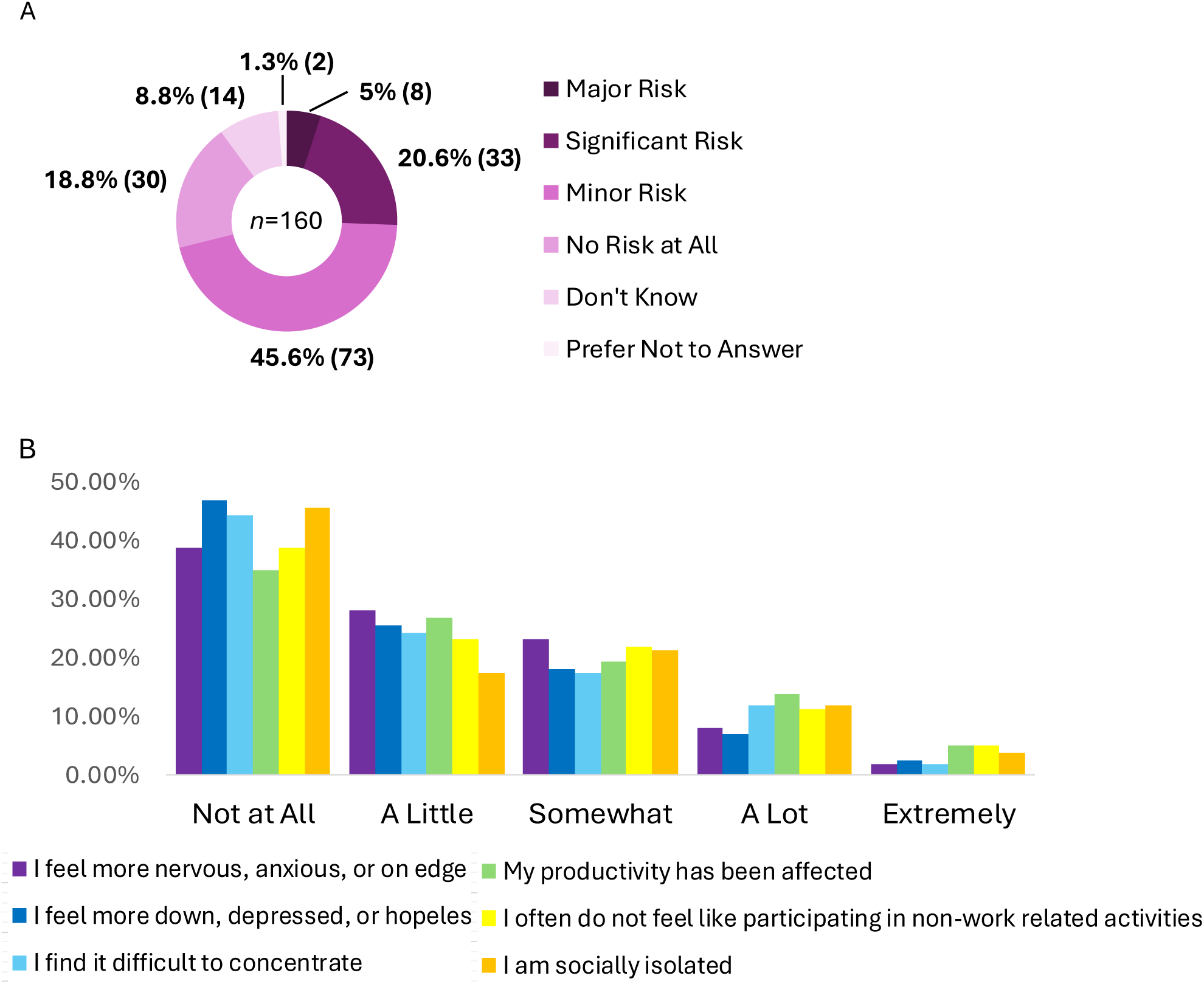
The impact of the COVID-19 pandemic on postdocs’ mental health and wellbeing. **A**) Overall percentages of each answer to the question “To what extent do you think the coronavirus (COVID-19) pandemic poses a risk to your mental health?”. **B**) Graphic representation of how the COVID-19 pandemic has impacted each listed feeling.

Next, postdoctoral researchers were asked in which way the pandemic affected their wellbeing in regard to certain mental states and moods associated with psychological distress: feelings of anxiety or depression, productivity levels, and levels of sociality. In line with the perception of the pandemics’ risks, the majority of respondents (62-73% felt that the pandemic had no to little impact on any of the proposed psychological distresses (**Figure 4B and Figure S2**). However, a notable percentage of the respondents (9-19%%) declared that the pandemic still affected a lot or extremely some aspects of their life, with the highest impacts being productivity (18.75%) and social interactions (participating in non-work related activities: 16.25%, and feeling socially isolated: 15.63%). Among Asian respondents, we noted that the effect of the pandemic on sociability is even more pronounced as over 25% identified that the pandemic had “extremely” or “a lot’ impacted their feeling of isolation and their interest in participating in social activities, with other ethnicities falling between 5.56% and 16.67% (**Supplementary Data**).

#### Activities to improve mental health

It is well established that socialization, physical activity, and music improve mental health^30–36^. To assess the level of awareness and self-care of postdoctoral researchers, we asked them if they practice any of the listed activities to improve their mental health (**Table 4**). Over 2/3 of the respondents choose to meet friends (66.25%) to counteract stress and support mental health. Doing physical activity and yoga (63.13%), playing or listening to music (61.25%), and baking (53.75%) also appear to be the preferred choices by more than half of the respondents. Reading (45%), listening to podcasts (43.75%), and having a good sleep routine (41.88%) are also activities appreciated by over 2/5 of the postdoctoral researchers, while fewer use meditation (25%) and electronic games (21.25%) to support their mental health. A small but alarming 9.38% declared feeling a general lack of motivation preventing them to do any activity. These results reveal a solid awareness among postdoctoral researchers about the importance of work-independent activities for the mental health and general well-being.

**Table 4.** Percentage of respondents indicating the activity/hobby they conduct to support their general wellbeing; *n*=160

| Activity to support mental health |  |
| --- | --- |
| Meet friends | 66.25% |
| Physical activity, yoga | 63.13% |
| Music including listening to music or playing an instrument | 61.25% |
| Cook, bake | 53.75% |
| Read | 45.00% |
| Listen to podcasts, audiobooks | 43.75% |
| Good sleep routine | 41.88% |
| Meditation, mindfulness | 25.00% |
| Electronic games | 21.25% |
| None. I feel a general lack of motivation. | 9.38% |
| Other | 3.13% |

## DISCUSSION

With this survey we gathered important data that pave the way for future studies analyzing the mental health and well-being of postdoctoral researchers in academia. The demographic analysis revealed notable disparities in immigration status and ethnicity distributions. Specifically, a substantial proportion of respondents are non-permanent visa holders (74%), which corresponds to the overall WCM population (75%) (**Figure S3**), but not as much to the nation-wide distribution of domestic versus immigrant postdoctoral researchers that sees 44.6% of US citizens or permanent residents and 55.4% of temporary visa holders. A similar disparity between survey, WCM and national demographics can be observed for the gender distribution. Notably, the respondents were majority female. This gender ratio is slightly different from the gender distribution of WCM where in 2023 49% of postdoctoral researchers were male, 46% females and 5% non-disclosed (which is in line with the survey’s gender non-conform/undisclosed group) (**Figure S3**). However, on the national level, the gender distribution shifts even more compared to our survey, with only 28.3% of female versus 71.7% of male postdoctoral researchers at federally funded research insitutions and development centers^37–39^. The racial groups are more difficult to compare as ethnicities are often stratified differently. Our survey comprised 11% UriM, slightly less that 17% present at WCM, 29.5% Asian, and 59.5% identifying as belonging to ethnicities other than UriM or Asian, while at WCM the population of non-UriM postdoctoral researchers is 79%. These demographic differences in the survey are reflected in variations observed in certain responses when stratified by immigration status or ethnicity, which will be discussed in detail below.

### The inherent nature of researchers as (im)migrants

Academic researchers are often expected to relocate – across states, countries, and continents – to advance their careers. However, the distress caused by such long-distance moves is frequently underestimated. Relocating away from an established social network is inherently disruptive, and building a new one can be particularly challenging, especially in a large city and during formative ages for marriage/partnership, raising family, and care for aging parents. Our survey revealed that, after salary, work-life balance is the second most frequently cited factor influencing mental well-being (**Figure 2A**), and approximately one in six respondents reported feeling socially isolated or avoiding social events as a lingering consequence of the COVID-19 pandemic (**Figure 3B**). Similarly, social bonding activities (i.e., meeting friends) emerged as the most preferred strategy for supporting mental health and wellbeing, with two-thirds of participants selecting this option (**Table 4**). These answers all point towards the importance of having a strong social network, while the high pressure, long hours, and relocations inherent to this job, make it extremely challenging to devolve sufficient time for fostering social bonds and community.

Moving to a foreign country often requires a nonimmigrant work visa, which not only can be a nerve-wrecking and costly procedure but is often associated to additional stressors such as visa-related insecurity and instability. It is, therefore, not surprising that visa-related issues and the dependency between performance and visa status are perceived as heavily influencing visa-holders’ mental health (**Figure 2D, Figure S1 and Supplementary data**). Furthermore, issues related to immigration status have changed drastically in the US due to results of the 2024 presidential election. The current federal administration has revoked visas for over 1,000 students and recent graduates across more than 170 institutions in at least 40 states^40^. This crackdown has targeted individuals on F-1 academic and J-1 exchange visas with visa revocations occurring in many cases without communication of explanation or due process, having higher education associations asking for clarity from Homeland Security and the State Department^41^. These actions have immediate consequences on researcher’s careers and it is plausible that this type of sudden change and instability will have a serious impact on mental health as well.

### The Perception and Effect of Micro- and Macro-aggressions in Academic Workspaces

Our results found that more than half of respondents reported experiencing micro- and macro-aggressions highlighting the prevalence of this issue within academic institutions. However, a significant portion of those affected (40%) indicated that microaggressions did not have a substantial impact on their mental health, while 25% experienced little or moderate effects, and 10% were affected extensively. Similar percentages are true for macro-aggressions with 43% reporting not being much affected by them, 24% and 27% feeling respectively little and moderately affected, and only 6% felt extensively affected. One possible explanation for these unanticipated responses is that postdoctoral researchers may have developed coping mechanisms to mitigate the psychological impact of such experiences. Alternatively, the continuous and repeated exposure to micro- and macro-aggressions over time could result in a sense of desensitization, reducing the perceived severity of their impact on respondents’ mental health and potentially masking long-term consequences for mental health and well-being. Future research should explore the effects of microaggressions and the dynamic between resilience, coping strategies, and organizational culture in mitigating or exacerbating these impacts.

### The Persisting Impact of the COVID Pandemic

The COVID-19 pandemic has had a profound global impact and has been the focus of numerous studies, including those examining its effects on early-career researchers such as PhD students and postdoctoral researchers^19,42^. Motivated by these studies, we wanted to investigate whether, nearly four years after the outbreak, postdoctoral researchers at WCM—who experienced at least part of their doctoral training during this unprecedented period—still feel its effects. During the initial surge of the COVID-19 pandemic in New York, WCM implemented a brief, behavioral skills-based intervention program, “CopeNYP”, to address the immediate mental health needs of the employees of the hospital and medical school, including postdoctoral researchers from March 2020 to April 2021^43^. The data published from this program revealed a close association between the average number of CopeNYP sessions and average number of COVID-19 hospitalization over a 14 day period; specifically more employees of WCM sought out services to address their mental health as COVID hospitalizations were increasing in the spring of 2020 and 2021 and decreased their usage of these services in the summer and fall of 2020 as COVID hospitalizations decreased^43^. Given that maximum COVID hospitalizations in NYC were 80% lower between December 2023 to January 2024, when our survey was conducted, compared to the maximum hospitalizations that occurred during the CopeNYP program; our findings that postdoctoral researchers have low perception of risk of the COVID-19 pandemic impacts to their mental health is consistent with CopeNYP findings^44^. The changes observed in our study and the CopeNYP study may also reflect behavioral and societal shifts over time to COVID, which include the desire for widespread adoption of masking, improved understanding of COVID-19 transmission, and the availability of vaccines, which collectively could reduce the perceived threat of the pandemic. Furthermore, the timing of the studies relative to the initial lockdown may have influenced perceptions; during the early phases of the pandemic in 2020 and 2021, uncertainty and fear were heightened, whereas by late 2023, many had adapted to the “new normal,” further lessening the psychological toll.

### The impact of the current Administration on Research Institutions

The results of our survey highlight the mental health crisis among postdoctoral researchers, with high levels of anxiety and psychological distress reported in early 2024. These findings are particularly concerning in light of recent federal actions that exacerbate those very aspects that our survey revealed as key stressors, such as low salaries, stalled professional progress and a pervasive sense of instability. In particular, postdoctoral researchers now face heightened uncertainty due to NIH communication freezes, termination of previously awarded grants, disruption of planned study sections for grant evaluations, hiring freezes at academic institutions and proposed funding cuts to the NIH^45–47^. Such systemic disruptions not only threaten the stability of research careers but also strip away the chance to improve institutional support; as these institutions face existential threats, mental health and well-being are unlikely to become a priority during times of crisis. The culmination of these policy changes with an already vulnerable population signals a need for revisioning institutional interventions aimed at both protecting research infrastructure and supporting the well-being of its research community including its most vulnerable members.

### Intervention Strategies and Call to Action

To mitigate the identified challenges, we propose several concrete actions. First – despite understanding that the aforementioned current political situation represents a new, additional obstacle –raising postdoc salaries to reflect the cost of living in NYC would alleviate financial stress, a key driver of mental health stressors identified in this study. While the NIH sets minimum stipend levels for postdoctoral researchers funded through its grants, principal investigators (PIs) often have flexibility to supplement these salaries through other funding sources. However, this flexibility is not universally embraced or supported across academic institutions. Therefore, greater alignment and clarity between academic institutions and the NIH is necessary to reconcile this disparity and encourage PIs to provide more competitive compensation, ensuring that financial constraints do not detract from postdoctoral researchers’ well-being or productivity.

Second, the establishment and empowerment of postdoc unions could enhance job security and address workplace discrimination and abuse, providing postdoctoral researchers with a stronger voice in advocating for equitable treatment which is especially important in the current political climate. Expanding access to mental health resources to include free or subsidized psychological and psychiatric care would also offer critical support for postdoctoral researchers experiencing burnout, anxiety, or depression. These efforts should be accompanied by clear communication and awareness initiatives within the postdoctoral community to ensure their effective use. Additionally, academic institutions should invest in comprehensive career training programs to prepare postdoctoral researchers for diverse career paths, reducing the uncertainty surrounding their professional futures whether it be in academia or industry. Family support policies, including affordable childcare and paid leave for familial responsibilities, would further ease the burdens faced by postdoctoral researchers with caregiving duties. Third, fostering a sense of community and recognition through postdoc associations, funded social activities, and cross-departmental initiatives could improve social support and reduce isolation as well as expanding their professional network and fostering collaborations, which are essential for career progression. Leadership at the institutional and departmental levels must also play an active role in recognizing and valuing postdoctoral researchers, celebrating their contributions through awards, public acknowledgments, and integration into decision making processes. Finally, this study calls for heightened awareness and action to address the postdoc mental health crisis. Just as the mental health challenges of graduate students have received increased attention in recent years, the struggles of postdoctoral researchers should no longer be overlooked, and academic institutions and NIH should collaborate to implement lasting solutions to these problems.

### LIMITATIONS AND FUTURE DIRECTIONS

Our survey was designed to be a pilot study that begins to broadly describe the mental health situation of postdoctoral researchers; specifically, researchers who live in a large metropolitan city like New York City and work at a major medical research institution like WCM, an environment that allows for examination of unique struggles that are present in an understudied population of postdoctoral researchers^9^. To that end, we believe the sample size of our study is appropriate considering the size of the postdoctoral community at WCM; to ensure 85% power to detect an effect of *f=0.5* (alpha probability = 0.05), our survey needed a sample size of 145 responders. In line with our power analysis; our survey has 160 respondents and there are estimated to be 478 postdoctoral researchers at WCM^25^, resulting in a respondence rate of 33%. Based on this number, we decided to perform a qualitative analysis, especially considering that when stratifying by gender, ethnicity, and immigration status, certain subgroups resulted in a very small sample size (especially for gender non-conformed individuals, permanent residents, and UriM). Especially the identity of ethnicity would benefit from more granular categorizations, including more ethnicities to form a better picture of the true diversity. A potential limitation of our study is the possibility of self-selection bias among survey respondents, as individuals who are unhappy with their postdoctoral experiences may have been more likely to participate, potentially skewing the representativeness of the findings.

In the future, we hope to build on this study and conduct the survey again both at WCM and on a larger scale across the city, region, or nation, which would allow for a better representation of the national or even world-wide situation of postdoctoral researchers. Additional personal information such as age, country of origin, family status and living situation – single, married, co-living, children – as well as career-related details like years of postdoc, field of study, and average work hours are important factors that will be included in the future survey, as they contribute to psychological distress and mental disorders.

## CONCLUSIONS

Our study highlights significant challenges impacting the mental health and wellness of postdoctoral researchers, particularly those related to the demands of academic life and the complexities of living in a major metropolitan area. These findings underscore the pressing need for targeted action to address the unique stressors faced by this community, including those influenced by gender, ethnicity, and citizenship. The negative outlook on career progression and the difficulty transitioning to independent academic positions further emphasize the urgency of these issues. In line with past research, our results call for the establishment and expansion of mental health and career development resources for postdoctoral researchers. Institutions should leverage these insights to enhance support through updated policies, expanded resources within Postdoctoral Offices or Associations, and a cultural shift in academia to foster a more supportive environment for postdoctoral researchers.

## METHODS

### Survey Design

This exploratory survey was designed to capture the experiences and demographic information of postdoctoral associates at WCM. The survey was conducted over 2 months between December 2023 and January 2024 by the Weill Cornell Postdoctoral Association (PDA). The current survey includes validated measures from the Patient Health Questionnaire (PHQ-9) and General Anxiety Disorder (GAD-7) for mood symptoms in the General Mental Wellbeing sections^22,23^. The survey also gathered Information regarding depression, anxiety, and burnout related symptoms. At the end of the survey, the participants had a summary of mental health resources available both at WCM and in the community, including the National Suicide Prevention Lifeline, the NYC Sexual Violence Helpline, the New York State Domestic & Sexual Violence Hotline, and the National Drug Helpline.

### Data analysis

A total of 198 researchers currently holding postdoctoral positions at WCM in New York City, NY, submitted responses via the Qualtrics™ platform. After filtering for completeness, 160 valid responses were retained for further analysis using Qualtrics and Microsoft Excel. Graphical representations were generated using Microsoft Excel.

## Data Availability

All data produced in the present study are available upon reasonable request to the authors

## Notes

### Competing Interest Statement

The authors have declared no competing interest.

### Funding Statement

This study did not receive any funding

### Author Declarations

IRB of Weill Cornell Medical College gave ethical approval for this work.

## REFERENCES

1. Ahmed, M.Z., Plotkin, D., Qiu, B.-L., and Kawahara, A.Y. (2015). Postdocs in Science: A Comparison between China and the United States. BioScience 65, 1088–1095. 10.1093/biosci/biv125.

2. Levey, G.S., Sherman, C.R., Gentile, N.O., Hough, L.J., Dial, T.H., and Jolly, P. (1988). Postdoctoral research training of full-time faculty in academic departments of medicine. Ann Intern Med 109, 414–418. 10.7326/0003-4819-109-5-414.

3. Institute of, M., Committee on National Needs for, B., and Behavioral Research, P. (1984). The National Academies Collection: Reports funded by National Institutes of Health. In The Career Achievements of NIH Predoctoral Trainees and Fellows, P.E. Coggeshall, and P.W. Brown, eds. (National Academies Press (US) Copyright © National Academy of Sciences. All rights reserved.). 10.17226/19375.

4. Committee on Science, E., Public, P., Policy, Global, A., National Academy of, S., National Academy of, E., and Institute of, M. (2014). The National Academies Collection: Reports funded by National Institutes of Health. In The Arc of the Academic Research Career: Issues and Implications for U.S. Science and Engineering Leadership: Summary of a Workshop, (National Academies Press (US) Copyright 2014 by the National Academy of Sciences. All rights reserved.). 10.17226/18627.

5. Lu, J., Velten, B., Klaus, B., Ramm, M., Huber, W., and Coulthard-Graf, R. (2023). The changing career paths of PhDs and postdocs trained at EMBL. Elife 12. 10.7554/eLife.78706.

6. Postdocs in crisis: science cannot risk losing the next generation. (2020). Nature 585, 160. 10.1038/d41586-020-02541-9.

7. Woolston, C. (2022). Lab leaders wrestle with paucity of postdocs. Nature. 10.1038/d41586-022-02781-x.

8. Drosatos, K., and Fousteri, G. (2022). Generation LWBS: introducing life-work balance in science. Nat Cardiovasc Res 1, 1107–1108. 10.1038/s44161-022-00165-y.

9. Sainburg, T. (2023). American postdoctoral salaries do not account for growing disparities in cost of living. Research Policy 52, 104714. 10.1016/j.respol.2022.104714.

10. Kahn, S., and Ginther, D.K. (2017). The impact of postdoctoral training on early careers in biomedicine. Nat Biotechnol 35, 90–94. 10.1038/nbt.3766.

11. Elias, M.H., Sompiyachoke, K., Fernández, F.M., and Kamerlin, S.C.L. (2024). The ineligibility barrier for international researchers in US academia. EMBO Rep 25, 457–458. 10.1038/s44319-023-00053-x.

12. Woolston, C. (2020). Uncertain prospects for postdoctoral researchers. Nature 588, 181–184. 10.1038/d41586-020-03381-3.

13. Woolston, C. (2020). Postdocs under pressure: ’Can I even do this any more?’. Nature 587, 689–692. 10.1038/d41586-020-03235-y.

14. Culpepper, D., Reed, A.M., Enekwe, B., Carter-Veale, W., LaCourse, W.R., McDermott, P., and Cresiski, R.H. (2021). A New Effort to Diversify Faculty: Postdoc-to-Tenure Track Conversion Models. Front Psychol 12, 733995. 10.3389/fpsyg.2021.733995.

15. Arnold, C. (2014). The stressed-out postdoc. Science 345, 594. 10.1126/science.345.6196.594.

16. Dorsey, E.R., Jarjoura, D., and Rutecki, G.W. (2003). Influence of Controllable Lifestyle on Recent Trends in Specialty Choice by US Medical Students. JAMA 290, 1173–1178. 10.1001/jama.290.9.1173.

17. Pitt, R.N., Taskin Alp, Y., and Shell, I.A. (2021). The Mental Health Consequences of Work-Life and Life-Work Conflicts for STEM Postdoctoral Trainees. Front Psychol 12, 750490. 10.3389/fpsyg.2021.750490.

18. Afonja, S., Salmon, D.G., Quailey, S.I., and Lambert, W.M. (2021). Postdocs’ advice on pursuing a research career in academia: A qualitative analysis of free-text survey responses. PLoS One 16, e0250662. 10.1371/journal.pone.0250662.

19. Morin, A., Helling, B.A., Krishnan, S., Risner, L.E., Walker, N.D., and Schwartz, N.B. (2022). Surveying the experience of postdocs in the United States before and during the COVID-19 pandemic. eLife 11, e75705. 10.7554/eLife.75705.

20. Feld, L.D., Sarkar, M., Au, J.S., Flemming, J.A., Gripshover, J., Kardashian, A., Muir, A.J., Nephew, L., Orloff, S.L., Terrault, N., et al. (2023). Parental leave, childcare policies, and workplace bias for hepatology professionals: A national survey. Hepatol Commun 7. 10.1097/hc9.0000000000000214.

21. Demarinis, S. (2020). Loneliness at epidemic levels in America. Explore (NY) 16, 278–279. 10.1016/j.explore.2020.06.008.

22. Kroenke, K., Spitzer, R.L., and Williams, J.B. (2001). The PHQ-9: validity of a brief depression severity measure. J Gen Intern Med 16, 606–613. 10.1046/j.1525-1497.2001.016009606.x.

23. Spitzer, R.L., Kroenke, K., Williams, J.B., and Löwe, B. (2006). A brief measure for assessing generalized anxiety disorder: the GAD-7. Arch Intern Med 166, 1092–1097. 10.1001/archinte.166.10.1092.

24. Ko, M., and Ton, H. (2020). The Not Underrepresented Minorities: Asian Americans, Diversity, and Admissions. Acad Med 95, 184–189. 10.1097/acm.0000000000003019.

25. . https://postdocs.weill.cornell.edu/sites/default/files/2023_cngls-12_004_1.pdf.

26. Way, U. https://unitedwaynyc.org/true-cost-of-living/.

27. Torino, G.C., Rivera, D.P., Capodilupo, C.M., Nadal, K.L., and Sue, D.W. (2018). Microaggression Theory: What the Future Holds. In Microaggression Theory, pp. 307–328. 10.1002/9781119466642.ch19.

28. Pérez Huber, L., and Solorzano, D.G. (2015). Racial microaggressions as a tool for critical race research. Race Ethnicity and Education 18, 297–320. 10.1080/13613324.2014.994173.

29. Morin, A., Helling, B.A., Krishnan, S., Risner, L.E., Walker, N.D., and Schwartz, N.B. (2022). Surveying the experience of postdocs in the United States before and during the COVID-19 pandemic. Elife 11. 10.7554/eLife.75705.

30. Solomonov, N. (2023). Improving social reward responsivity and social connectedness in psychotherapies for late-life depression: Engage & Connect as an example. Psychiatry Research 329, 115469. 10.1016/j.psychres.2023.115469.

31. Solomonov, N., Victoria, L.W., Lyons, K., Phan, D.K., Alexopoulos, G.S., Gunning, F.M., and Flückiger, C. (2023). Social reward processing in depressed and healthy individuals across the lifespan: A systematic review and a preliminary coordinate-based meta-analysis of fMRI studies. Behavioural Brain Research 454, 114632. 10.1016/j.bbr.2023.114632.

32. Umberson, D., Crosnoe, R., and Reczek, C. (2010). Social Relationships and Health Behavior Across the Life Course. Annual Review of Sociology 36, 139–157. 10.1146/annurev-soc-070308-120011.

33. Calhoun, C.D., Stone, K.J., Cobb, A.R., Patterson, M.W., Danielson, C.K., and Bendezú, J.J. (2022). The Role of Social Support in Coping with Psychological Trauma: An Integrated Biopsychosocial Model for Posttraumatic Stress Recovery. Psychiatric Quarterly 93, 949–970. 10.1007/s11126-022-10003-w.

34. Cleary, J.L., Fang, Y., Zahodne, L.B., Bohnert, A.S.B., Burmeister, M., and Sen, S. (2023). Polygenic Risk and Social Support in Predicting Depression Under Stress. American Journal of Psychiatry 180, 139–145. 10.1176/appi.ajp.21111100.

35. Guay, S., Nachar, N., Lavoie, M.E., Marchand, A., and O’Connor, K.P. (2017). The buffering power of overt socially supportive and unsupportive behaviors from the significant other on posttraumatic stress disorder individuals’ emotional state. Anxiety, Stress, & Coping 30, 52–65. 10.1080/10615806.2016.1194400.

36. Løseth, G.E., Eikemo, M., Trøstheim, M., Meier, I.M., Bjørnstad, H., Asratian, A., Pazmandi, C., Tangen, V.W., Heilig, M., and Leknes, S. (2022). Stress recovery with social support: A dyadic stress and support task. Psychoneuroendocrinology 146, 105949. 10.1016/j.psyneuen.2022.105949.

37. Ceci, S.J., and Williams, W.M. (2011). Understanding current causes of women’s underrepresentation in science. Proceedings of the National Academy of Sciences 108, 3157–3162. doi:10.1073/pnas.1014871108.

38. Larivière, V., Ni, C., Gingras, Y., Cronin, B., and Sugimoto, C.R. (2013). Bibliometrics: Global gender disparities in science. Nature 504, 211–213. 10.1038/504211a.

39. NC. https://ncses.nsf.gov/surveys/ffrdc-postdocs/2023#analysis.

40. Caroll Alvarado, J.H.a.A.M. https://www.cnn.com/2025/04/17/us/university-international-student-visas-revoked/index.html.

41. Mitchell, T. https://www.acenet.edu/Documents/Letter-State-Visa-Revocations-040425.pdf.

42. Hall, S. (2024). How PhD students and other academics are fighting the mental-health crisis in science. Nature 631, 496–498. 10.1038/d41586-024-02225-8.

43. Kanellopoulos, D., Solomonov, N., Ritholtz, S., Wilkins, V., Goldman, R., Schier, M., Oberlin, L., Bueno-Castellano, C., Dargis, M., Cherestal, S., and Gunning, F. (2021). The CopeNYP program: A model for brief treatment of psychological distress among healthcare workers and hospital staff. General Hospital Psychiatry 73, 24–29. 10.1016/j.genhosppsych.2021.09.002.

44. COVID, N. https://coronavirus.health.ny.gov/daily-hospitalization-summary.

45. Meredith Wadman, J.K. https://www.science.org/content/article/trump-hits-nih-devastating-freezes-meetings-travel-communications-and-hiring.

46. Reardon, S. https://www.science.org/content/article/nih-freezes-all-research-grants-columbia-university.

47. Gabrielle Masson, D.I. https://www.fiercebiotech.com/biotech/nih-directed-not-tell-universities-about-new-grant-freezes-harvard-battle-escalates.

