## Supplementary Figures and Tables for "Assessing the Mental Health Crisis Among New York’s Postdoctoral Researchers"

**Table S1**: Extended version of Table 1, showing individual percentages for response categories: “never”, “rarely” , “frequently” and “always.”

| Symptoms | Never | Rarely | Sometimes | Frequently | Always | Total (*n*) |
|---|---|---|---|---|---|---|
| Trouble relaxing. | 2.50% | 13.13% | 36.25% | 35.63% | 12.50% | 160 |
| Feeling overwhelmed by responsibilities. | 5.00% | 15.63% | 33.13% | 25.62% | 20.63% | 160 |
| Feeling nervous. | 1.25% | 13.75% | 39.38% | 34.38% | 11.25% | 160 |
| Being depleted of energy. | 3.75% | 18.75% | 33.13% | 30.63% | 13.75% | 160 |
| Reduced self confidence. | 6.25% | 18.75% | 35.00% | 26.25% | 13.75% | 160 |
| Feeling down or sad. | 3.75% | 16.88% | 40.00% | 30.00% | 9.38% | 160 |
| Having a recurring feeling that something negative might happen. | 10.63% | 21.88% | 31.87% | 25.00% | 10.63% | 160 |
| Little interest in socializing. | 11.25% | 20.00% | 33.75% | 23.13% | 11.88% | 160 |
| Sleeping issues. | 10.00% | 23.13% | 31.87% | 21.25% | 13.75% | 160 |
| Trouble concentrating. | 5.00% | 24.38% | 38.13% | 21.25% | 11.25% | 160 |
| Fear of having disappointed important people in my life. | 16.25% | 23.13% | 28.13% | 18.75% | 13.75% | 160 |
| Becoming easily annoyed or irritable. | 5.63% | 27.50% | 39.38% | 18.75% | 8.75% | 160 |
| Poor appetite or overeating. | 16.25% | 23.75% | 32.50% | 20.00% | 7.50% | 160 |
| Slowed thinking process. | 6.25% | 30.00% | 36.88% | 18.13% | 8.75% | 160 |
| Feeling disconnected from your body. | 29.38% | 36.88% | 16.88% | 11.88% | 5.00% | 160 |
| Noticeable changes in movement speed or speech rate. | 37.50% | 36.25% | 16.25% | 6.88% | 3.13% | 160 |

**Figure S1** – Work-related factors that impact postdocs' mental health and general wellbeing stratified by immigration status: US-Citizens or Permanent Residents versus Visa Holders.

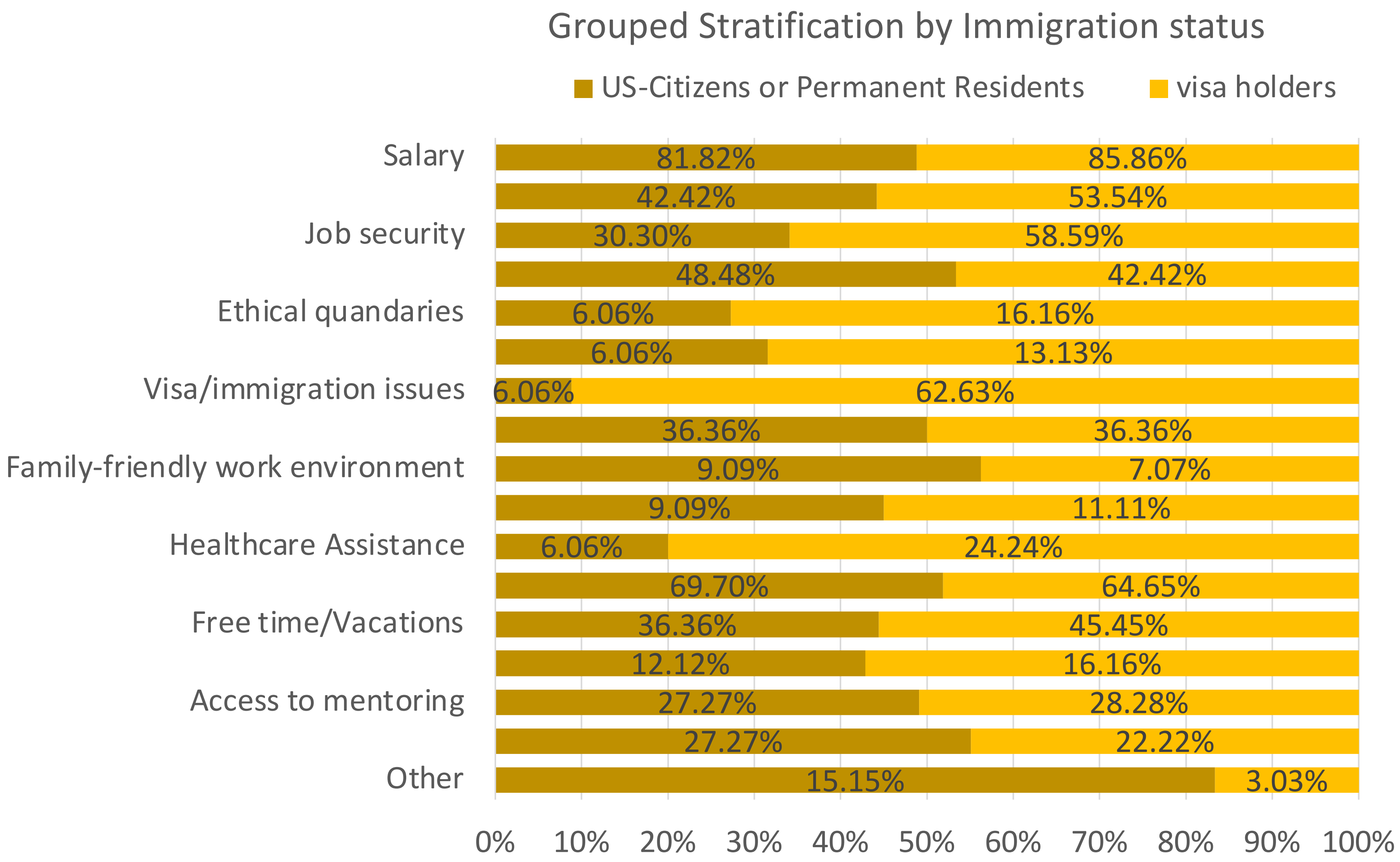

**Table S2** – Work-related factors that impact postdocs' mental health and general wellbeing stratified by immigration status: US-Citizens or Permanent Residents versus Visa Holders.

| **Work-related Stressor** | **Yes** | **No** | **Sometimes** | **Total (*n*)** |
|---|---|---|---|---|
| Relationship with PI | 43.10% | 31.30% | 25.60% | 160 |
| Relationship with co-workers | 31.30% | 45.00% | 23.80% | 160 |
| Pressure to work at optimum levels, all the time | 52.50% | 23.10% | 24.40% | 160 |
| Dependency between performance and our visa status | 40.00% | 43.10% | 16.90% | 160 |
| Lack of clarity regarding vacation policy | 33.10% | 50.00% | 16.90% | 160 |
| Insufficient support for dependents | 32.50% | 56.30% | 11.30% | 160 |
| Lack of career development opportunities | 47.50% | 24.40% | 28.10% | 160 |

**Figure S2** – Grouped answers to "In which way has COVID-19 pandemic affected your wellbeing?" displayed in Figure 4B.

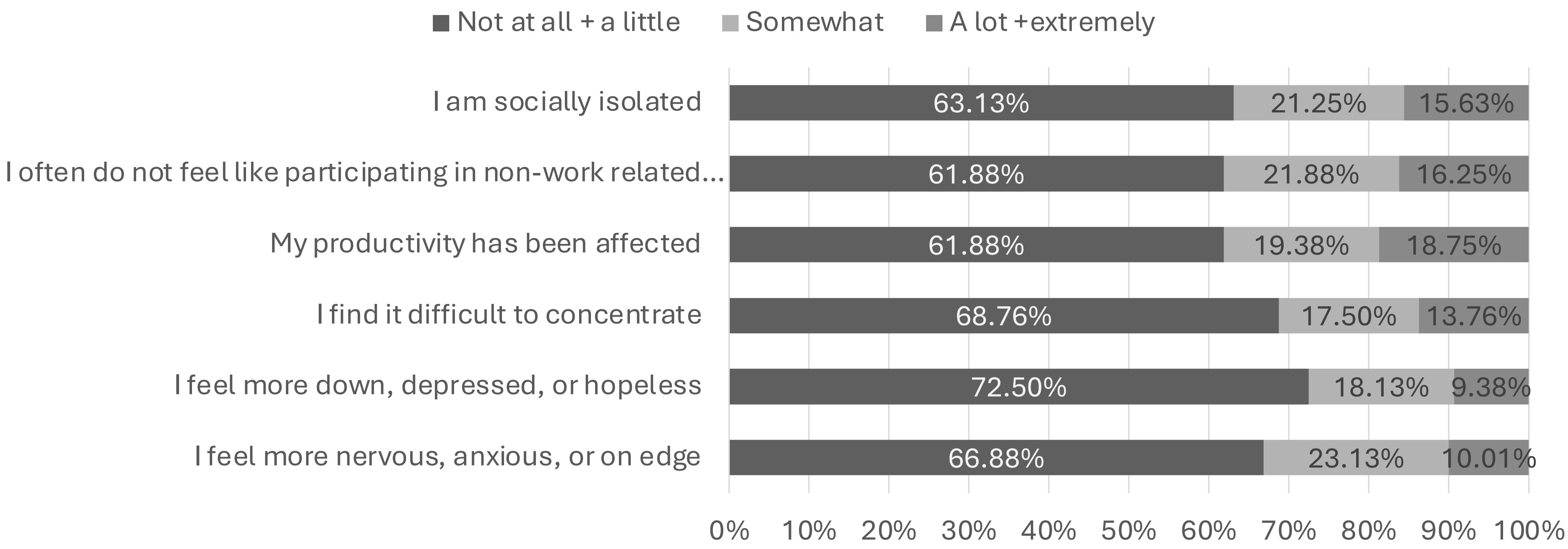

**Figure S3** – Demographics of Weill Cornell Medicine in 2023. *n* = 478. The total number of respondents for the Ethnicity is below 478 due to omitted responses.

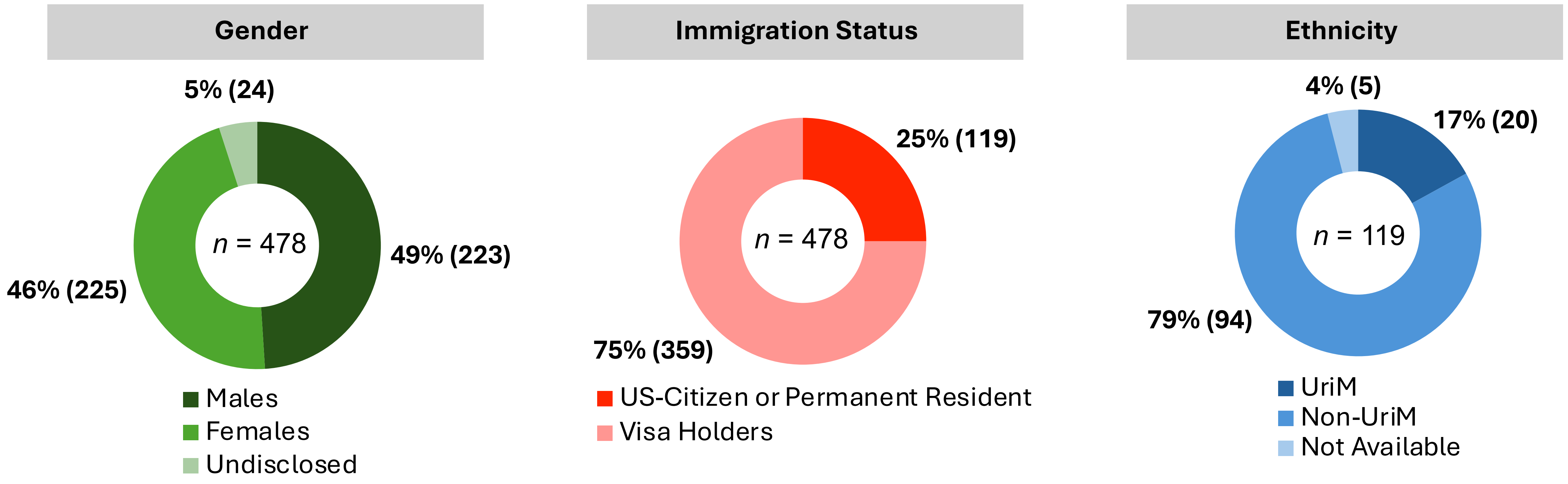
